# Selectively Augmented Decision Tree for Explainable Dementia Detection

**DOI:** 10.64898/2026.02.03.26345441

**Authors:** Firuz Kamalov, Fadi Thabtah, David Peebles, Anthony Ibrahim

## Abstract

Timely and accurate diagnosis of dementia remains a critical yet challenging task. Although machine learning (ML) techniques have shown considerable promise in dementia detection, their inherent complexity often results in opaque, “black-box” models that limit clinical acceptance and usability. In this paper, we propose a Selectively Augmented Decision Tree (SADT), an interpretable AI model specifically designed for dementia detection. SADT incorporates a structured three-phase pipeline consisting of feature selection, data balancing, and construction of a transparent decision tree classifier. We apply SADT to the OASIS dataset and evaluate it empirically, showing that SADT outperforms traditional ML benchmarks, validating its effectiveness. In addition to its superior performance, SADT also mirrors aspects of human decision-making in its sequential, rule-based prioritization of key features. This approach aligns with cognitive models of cue use and heuristic reasoning, making it not only clinically transparent but also psychologically aligned with how diagnostic decisions are often made in practice. SADT’s strong predictive performance and interpretability grounded in human reasoning facilitates explanation and human scrutiny, and has the potential to improve both clinical decision-making and trust in AI-assisted diagnosis.

## 1 Introduction

The accurate and timely diagnosis of dementia, particularly Alzheimer’s disease (AD), remains a critical global health challenge. Dementia is characterized by progressive cognitive decline, which significantly impacts patients’ quality of life and imposes substantial economic and emotional burdens on healthcare systems, patients, and care-givers [24]. Despite advances in diagnostic techniques, traditional clinical assessments and neuroimaging methods often encounter limitations in early and precise dementia detection, thus necessitating innovative methodologies to address these diagnostic challenges [18].

Artificial Intelligence (AI), especially machine learning (ML), has demonstrated considerable potential in enhancing the accuracy and efficiency of dementia diagnosis. AI methods enable automated analysis of complex data derived from medical imaging, clinical records, and cognitive assessments [30]. While ML models such as deep learning algorithms have achieved notable successes in identifying disease-specific patterns from large datasets, their inherent complexity often results in opaque decision-making processes. This “black-box” nature of AI models poses significant challenges, particularly in healthcare contexts, where interpretability and transparency are critical for clinical decision-making, patient communication, and ethical compliance [36].

Explainable AI (XAI) methods have consequently emerged as crucial tools for medical applications. XAI approaches address this interpretability gap by providing clarity and insight into the model’s decision-making processes. Techniques such as Local Interpretable Model-agnostic Explanations (LIME) [32], SHapley Additive exPlanations (SHAP) [25], and Gradient-based Saliency Maps [2] have garnered particular attention due to their capacity to generate localized and interpretable explanations. However, these methods typically offer indirect interpretations, highlighting the need for inherently interpretable models that enable direct scrutiny of their internal logic.

In this paper, we propose a novel explainable AI model specifically designed for dementia detection. Our approach leverages a dataset comprising clinical and anatomical measures derived from brain imaging data. The proposed methodology integrates three principal components: feature selection, data balancing, and interpretable model building. Effective feature selection enhances interpretability by reducing the complexity of the model. Data balancing addresses the class imbalance prevalent in dementia datasets, where positive instances constitute a minority. Finally, our interpretable model enables direct analysis of classification processes, thus surpassing the limitations of indirect interpretation methods like LIME.

Numerical experiments demonstrate that the proposed model achieves superior classification accuracy compared to existing state-of-the-art methods. This paper’s primary contributions include the design of an interpretable AI model and the improvement of dementia detection accuracy, thereby facilitating early diagnosis and promoting broader clinical adoption of transparent AI diagnostic tools.

In addition to its clinical value, the SADT model also raises interesting questions regarding the cognitive alignment of AI models. Decision trees may not only be interpretable in a formal sense but may also resonate with how expert clinicians and laypeople organize and apply diagnostic knowledge. Cognitive science research suggests that people often use rule-based or heuristic strategies, for example fast-and-frugal decision trees [15], to make efficient and intelligible judgments under uncertainty. By creating transparent, hierarchical decision boundaries, SADT may support diagnostic reasoning in ways that align with human categorization strategies and cue-based reasoning [7, 33]. This alignment with human cognitive decision making processes may engender greater trust and explainability in human-AI interaction, which would be a significant strength for clinical adoption.

The remainder of this paper is structured as follows. Section 2 provides a review of the relevant literature in dementia detection and existing XAI methodologies. Section 3 presents a detailed description of the proposed interpretable AI model. We outline the feature selection strategies, data balancing techniques, and the interpretability mechanisms employed. Section 4 reports the results of extensive numerical experiments that demonstrate the effectiveness of our approach against current state-of-the-art methods. Finally, Section 5 concludes the paper with a discussion of the key findings, their implications, and suggestions for future research directions.

## 2 Literature Review

Recent research has increasingly utilized non-imaging clinical data such as neuropsychological assessments, electronic health records (EHR), and genetic biomarkers for dementia detection. While traditional statistical methods remain relevant, modern ML techniques, particularly ensemble methods such as XGBoost and Random Forests, have proven superior in handling high-dimensional datasets and capturing complex interactions [10, 14].

Complementing the ensemble approaches, Thabtah and Peebles [37] introduced the Alzheimer’s Disease Class Rules (AD-CR) algorithm, an interpretable rule-based classifier that achieved 92.4% accuracy, 91.3% sensitivity, and 93.5% specificity on ADNI subsets. Similarly, AlShboul et al. [3] showed that classical models such as C4.5 and Naïve Bayes, trained on baseline Clinical Dementia Rating Sum-of-Boxes (CDR-SB) scores and demographics, reliably distinguished CN, MCI, and dementia groups, confirming that a single comprehensive cognitive instrument paired with lightweight ML can support rapid, non-invasive screening.

For example, Chun et al. [10] reported superior performance using XGBoost to predict conversion from mild cognitive impairment (MCI) to Alzheimer’s dementia (AUC = 0.85). They highlighted age, cognitive test scores, and APOE genotype as key predictors. Tang et al. [35] similarly employed Random Forests on extensive EHR data to predict Alzheimer’s years before diagnosis (ROC-AUC up to 0.81), identifying conditions such as hyperlipidemia – a condition genetically linked to Alzheimer’s – as an early predictive marker.

Further integrating genetic and clinical data, Gao et al. [14] utilized polygenic risk scores (PRS) alongside medical records within an XGBoost framework, achieving high accuracy (AUC = 0.88). They demonstrated the model’s interpretability through SHAP, highlighting genetic risk as crucial in older populations, while lifestyle factors were predominant in younger cohorts [14]. Additionally, language-based features derived from clinical text using NLP methods have shown promise in dementia sub-type classification, illustrating interpretability through clinically meaningful features [21].

Explainability remains central to non-imaging ML approaches. Jahan et al. [19] utilized multiple explainable AI (XAI) techniques (SHAP, LIME, partial dependence) to clarify ensemble predictions, whereas Cai et al. [9] implemented inherently interpretable Explainable Boosting Machines (EBMs) to transparently model dementia progression. Kubi and Nazir [22] further emphasized fairness in dementia predictions, ensuring demographic biases were mitigated via SHAP analyses. Collectively, these studies underscore the significance of age, genetic predisposition, cognitive status, and metabolic comorbidities as consistently important predictors, though generalization beyond specific cohorts remains a challenge [10, 14].

Neuroimaging has increasingly adopted convolutional neural networks (CNNs) for dementia classification due to their capacity to directly learn complex spatial features from MRI and PET images. High classification accuracies (*>*90%) in distinguishing Alzheimer’s disease (AD) from controls are frequently reported [20]. Böhle et al. [5] illustrated CNN interpretability via Layer-wise Relevance Propagation (LRP), producing heatmaps aligning closely with known AD pathology such as hippocampal atrophy. Similarly, Iizuka et al. [17] used Grad-CAM with CNNs on perfusion SPECT to identify dementia with Lewy bodies by accurately localizing the clinically recognized “cingulate island sign”.

Hybrid imaging approaches integrating clinical data also enhance interpretability. Lew et al. [23] combined MRI-derived CNN features with clinical information to accurately predict Alzheimer’s biomarker status, clarifying model decisions through logistic regression coefficients and Grad-CAM visualizations. Multimodal data fusion further extends interpretability, as demonstrated by Jahan et al.[19], who showed MRI dominance in distinguishing Alzheimer’s while cognitive assessments improved accuracy for early-stage cases. Advanced methods, including transformer-based and graph convolutional networks, are increasingly explored for their intrinsic interpretability potential [39].

Our review shows that both non-imaging and imaging ML methods effectively aid dementia detection. Their primary difference lies in data type and interpretability approaches. Non-imaging models leverage large datasets for early prediction using ensemble methods, whereas imaging models, particularly CNNs, achieve higher diagnostic precision by capturing subtle neuropathological changes [10, 20]. Explainability through SHAP, LIME, or visual heatmaps is common to both, though external validation and clinician-focused interpretation remain underexplored [27].

We find that several research gaps persist, notably regarding model generalization due to dataset homogeneity [27]. Limited prospective clinical validation further hinders practical deployment. XAI explanations also require rigorous validation, as current interpretability methods may sometimes provide misleading insights [38]. Additionally, dementia subtypes beyond Alzheimer’s remain understudied, representing a critical gap in current ML applications [17, 21].

Future research should thus focus on developing integrated multimodal predictive models capable of combining imaging, clinical, and longitudinal data within transparent frameworks [9, 19]. Further, embedding explainability into model training could improve interpretability and clinical acceptance, enhancing clinician-AI collaboration [27]. Addressing these gaps and opportunities will facilitate the transition of ML-based dementia detection from research settings to routine clinical practice.

## 3 Selectively Augmented Decision Tree (SADT)

The proposed dementia detection method SADT consists of a structured pipeline comprising three principal phases: (1) feature selection, (2) data balancing, and (3) construction of an interpretable classification model. These sequential stages are designed to enhance model interpretability, address inherent data imbalance, and ultimately enable transparent and accurate diagnosis of dementia. Figure 1 illustrates an overview of the pipeline.

**Fig. 1.**
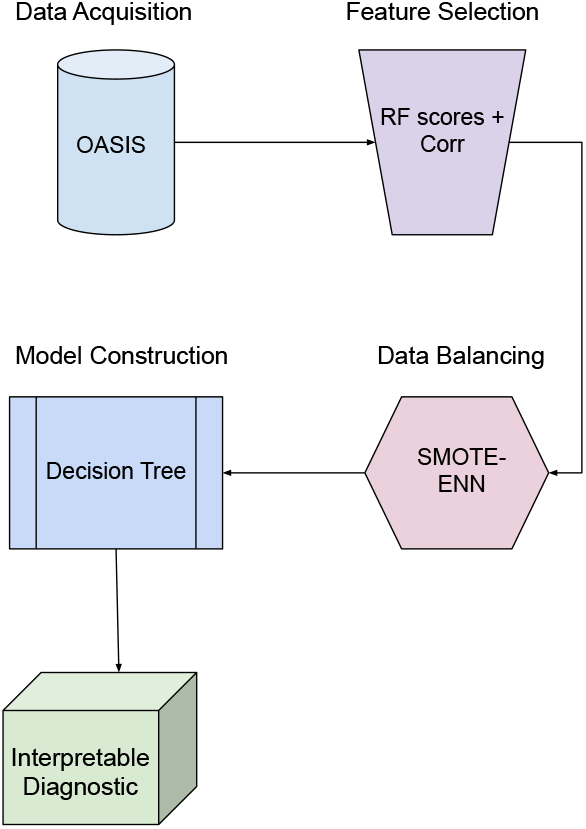
Selectively Augmented Decision Tree (SADT) pipeline.

### 3.1 Feature Selection

Feature selection is a crucial first step toward developing an interpretable model. Large feature sets can obscure meaningful relationships within data, complicate model interpretability, and may lead to suboptimal predictive performance due to redundancy and noise. To identify the most informative predictors, we apply a two-fold strategy based on Random Forest feature importance and correlation analysis.

Initially, a Random Forest classifier is trained to evaluate and rank feature importance based on the Gini impurity criterion. Features receiving low importance scores are considered less relevant and therefore excluded. Next, pairwise correlation analysis is conducted to identify and eliminate highly correlated features, thus further reducing multicollinearity and redundancy in the selected subset. Specifically, if two features exhibit high correlation, the less important feature (as determined by Random Forest rankings) is removed. This iterative process results in a refined subset of highly informative yet distinct predictors, significantly enhancing interpretability and analytical clarity.

The feature selection process can be understood not only in statistical terms, but also in terms of its cognitive relevance. In cognitive psychology, effective decision-making is often modeled as the application of selective attention to high-validity cues and the suppression of redundant or weakly informative features [7, 16]. Our approach using Random Forest feature importance followed by correlation filtering can be considered to be a computational analogue of this human strategy, prioritizing features with both high diagnosticity and minimal redundancy. This alignment with human cue selection processes may enhance the model’s psychological plausibility and strengthen its role in supporting human decision making.

### 3.2 Data Balancing

Dementia-related datasets frequently suffer from significant class imbalance, where negative instances (non-demented individuals) substantially outnumber positive cases (individuals diagnosed with dementia). Such imbalance can adversely affect the performance of classifiers, and result in biased predictions and reduced sensitivity in detecting minority class instances.

To mitigate this challenge, we employ the SMOTE-ENN (Synthetic Minority Over-sampling Technique combined with Edited Nearest Neighbors) algorithm [6]. SMOTE-ENN integrates the benefits of synthetic minority instance generation from SMOTE with data cleaning capabilities provided by the Edited Nearest Neighbors (ENN) approach. Specifically, SMOTE generates new synthetic instances by interpolating among existing positive instances, thereby augmenting the underrepresented class and balancing the data. Then, ENN identifies and removes noisy or overlapping instances which results in a cleaner and more distinctly separable class distribution.

Compared to conventional methods such as SMOTE alone, SMOTE-ENN offers several advantages. It reduces the likelihood of overfitting caused by synthetic data, enhances classifier generalization by eliminating borderline or ambiguous samples, and typically achieves superior classification performance, especially in highly imbalanced medical datasets. Consequently, applying SMOTE-ENN contributes directly to improved accuracy and reliability in dementia detection outcomes [11].

### 3.3 Interpretable Model Construction

In the final stage of the pipeline, an interpretable classification model is constructed using the preprocessed, balanced dataset. Given the clinical requirement for transparency and ease of interpretation, we adopt decision trees as our model of choice.

Decision trees inherently provide an easily understandable, rule-based structure that explicitly illustrates the decision-making pathway from input features to classification outcomes.

The interpretability of decision trees is particularly advantageous in clinical scenarios, where understanding the basis of diagnostic decisions is critical for clinical acceptance, patient communication, and ethical considerations. Moreover, the transparent, hierarchical structure of decision trees enables direct scrutiny of the classification logic, obviating the need for external post-hoc explainability methods.

The structural properties of the SADT decision tree align with cognitive models of human categorization. In particular, the tree’s simple, threshold-based rules resemble rule-plus-exception models and prototype-based categorizations [31], where diagnostic categories are formed by attending to key features and their combinations. In this case, MMSE functions as a primary cue while nWBV acts as a secondary criterion for borderline cases. Empirical studies have shown that clinicians often employ fast-and-frugal heuristics, and that such heuristic strategies, while efficient, can lead to systematic errors [4]. It highlights the need for models that align with human decision-making processes but avoid such errors. In addition, transparent, human-like justifications may result in increased explanation satisfaction in clinicians, boosting confidence in decisions [28]. This has important implications for AI decision support systems where explanations need to be not only technically correct but also convincing and cognitively satisfying to users.

### 3.4 Application to the OASIS Dataset

To illustrate the practical utility and efficacy of our proposed methodology, we apply the described pipeline to the Open Access Series of Imaging Studies (OASIS) dataset [26]. This dataset comprises demographic information, clinical assessments, and derived anatomical measurements obtained through brain imaging. The structured nature of the dataset allows for thorough assessment of the proposed pipeline, demonstrating the effectiveness of our approach.

#### 3.4.1 OASIS dataset

The OASIS dataset employed in our study consists of several demographic, clinical, and anatomical features obtained from 416 subjects. Our task is to predict the target variable Clinical Dementia Rating (CDR) based on various input features. The data is filtered to remove entries with missing values. The description of the input variables is given below.

##### Demographics

Age, Education (Educ), socioeconomic status (SES) [34]. Education codes correspond to the following levels of education: 1: less than high school grad., 2: high school grad., 3: some college, 4: college grad., 5: beyond college.

##### Clinical

Mini-Mental State Examination (MMSE) (Rubin et al., 1998), Clinical Dementia Rating (CDR; 0= nondemented; 0.5 – very mild dementia; 1 = mild dementia [29]. All participants with dementia (CDR*>*0) were diagnosed with probable AD.

##### Derived anatomic volumes

Estimated total intracranial volume (eTIV) (mm3), Atlas scaling factor (ASF) [8], Normalized whole brain volume (nWBV) [13].

The summary of the key statistics of the OASIS data is presented in Table 1.

**Table 1.** Summary of OASIS data.

|  | Age | Educ | SES | MMSE | eTIV | nWBV | ASF | CDR |
| --- | --- | --- | --- | --- | --- | --- | --- | --- |
| count | 436 | 235 | 216 | 235 | 436 | 436 | 436 | 235 |
| mean | 51.36 | 3.18 | 2.49 | 27.06 | 1481.92 | 0.79 | 1.2 | 0.29 |
| std | 25.27 | 1.31 | 1.12 | 3.7 | 158.74 | 0.06 | 0.13 | 0.38 |
| min | 18 | 1 | 1 | 14 | 1123 | 0.64 | 0.88 | 0 |
| 25% | 23 | 2 | 2 | 26 | 1367.75 | 0.74 | 1.11 | 0 |
| 50% | 54 | 3 | 2 | 29 | 1475.5 | 0.81 | 1.19 | 0 |
| 75% | 74 | 4 | 3 | 30 | 1579.25 | 0.84 | 1.28 | 0.5 |
| max | 96 | 5 | 5 | 30 | 1992 | 0.89 | 1.56 | 2 |

#### 3.4.2 Feature selection

To identify the relevant features in classifying CDR, we employ a combination of feature importance score derived from a fitted Random Forest model and pairwise correlation between the variables in the dataset. As shown in Figure 2, the Random Forest model found MMSE (importance: 0.29) and nWBV (importance: 0.19) to be the most influential features. The correlation analysis presented in Figure 3 also confirms that MMSE (-0.75) and nWBV (-0.50) have the strongest linear relationships with CDR. Thus, the converging evidence from feature importance and correlation analyses supports selecting MMSE and nWBV as the main predictors for CDR.

**Fig. 2.**
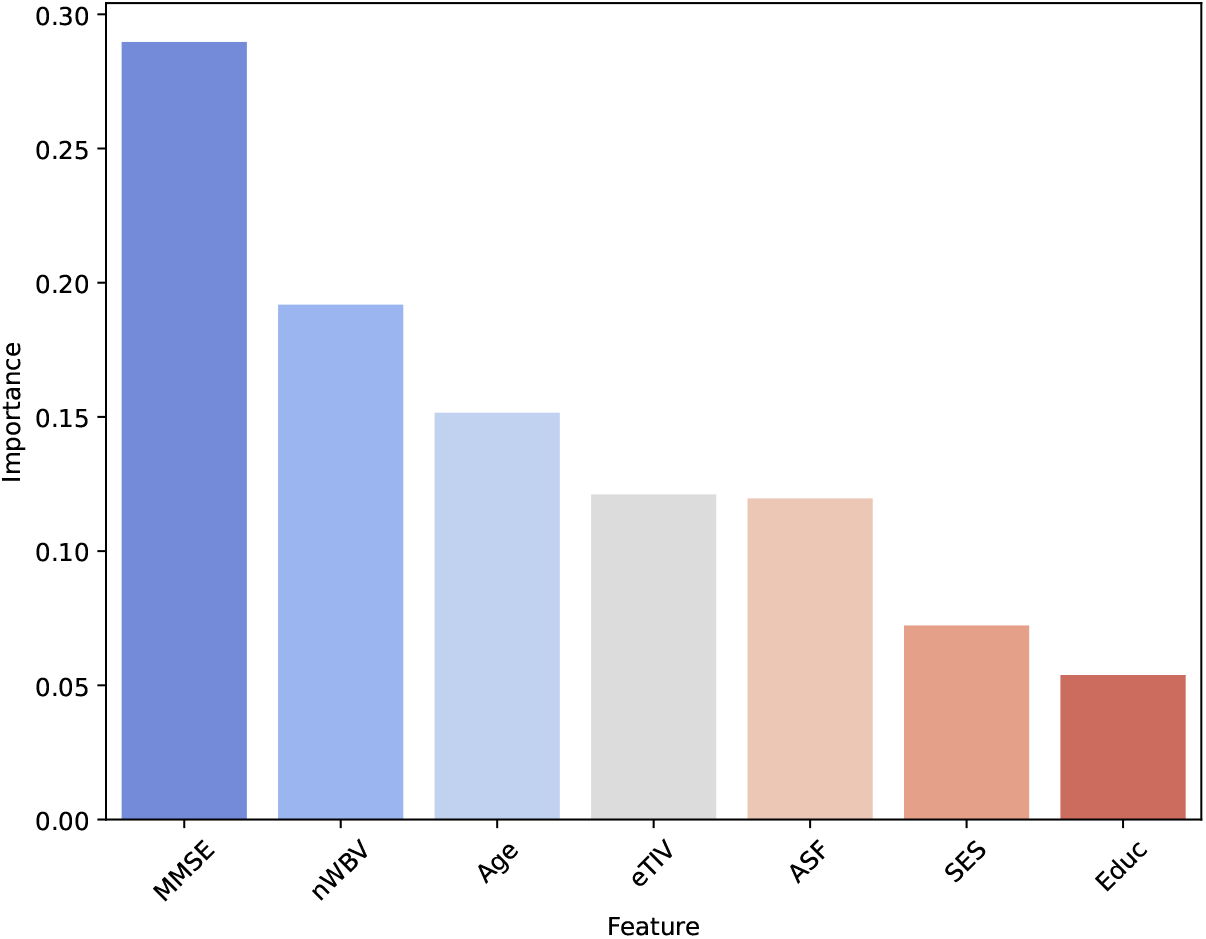
Random Forest feature importances.

**Fig. 3.**
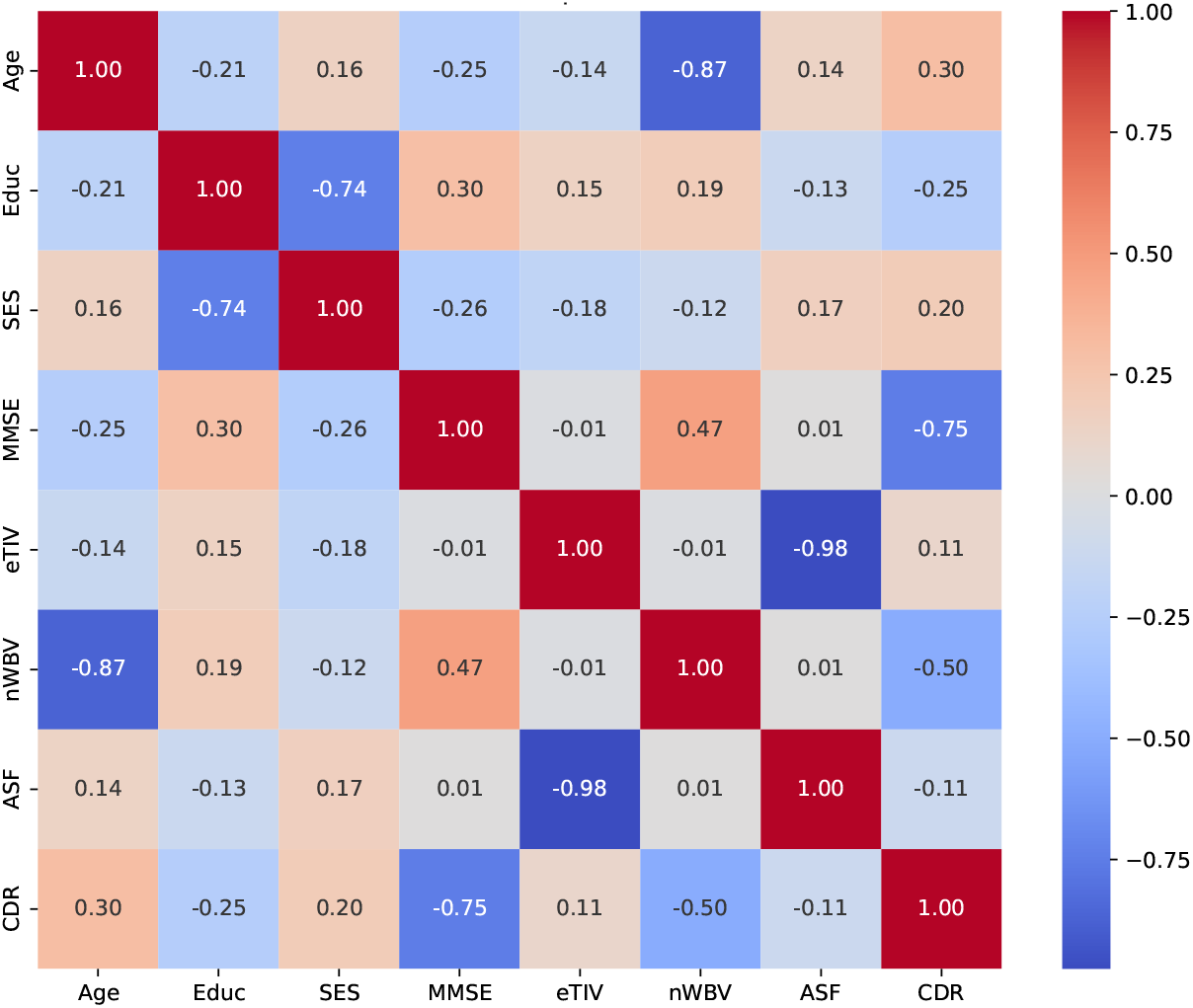
Pairwise correlation heatmap for the OASIS dataset.

Although Age ranks third in feature importance (0.15), it has a high negative correlation with nWBV (-0.87), indicating redundancy. Since nWBV already captures most of Age’s predictive information, focusing on MMSE and nWBV alone is sufficient. These two features provide a simple yet effective basis for predicting CDR.

#### 3.4.3 Data balancing

As shown in Figure 4, the distribution of CDR is skewed with the majority of instances labeled as 0. The SMOTE-ENN algorithm is applied to balance the data. In particular, the filtered dataset from the previous stage consisting of only MMSE and nWBV features is oversampled, to increase the number of the minority class instances to that of the majority class.

**Fig. 4.**
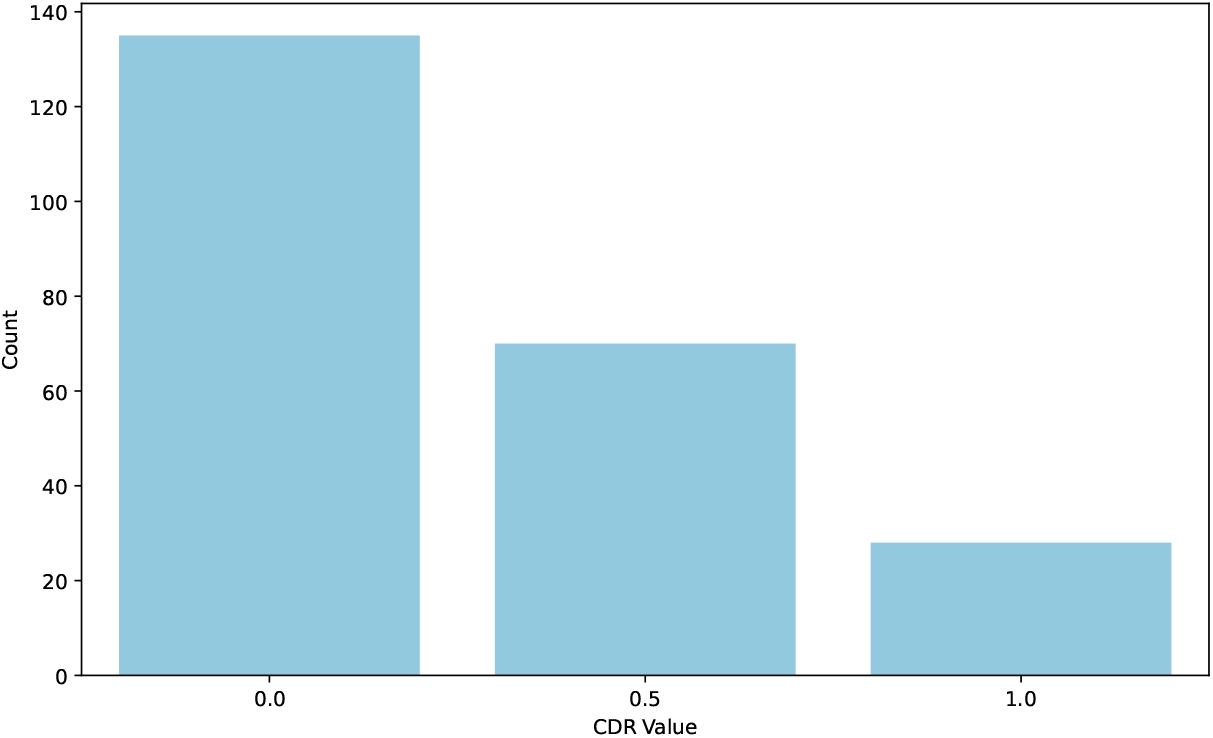
Distribution of CDR class labels.

#### 3.4.4 Explainable Model

We utilize the balanced dataset, comprising the selected features MMSE and nWBV, to construct an explainable model for dementia detection. Given the clinical need for interpretability, we chose a decision tree classifier for its transparency and straight-forward interpretability. Decision trees explicitly reveal decision logic, making them suitable for medical applications where understanding model predictions is critical.

The decision tree classifier is trained using the training subset of the balanced dataset. The derived decision tree is illustrated in Figure 5. At the root node, the tree evaluates the MMSE feature, applying a threshold value of approximately 27.57 to partition the dataset into two major groups. Subjects with MMSE values less than or equal to the threshold tend toward dementia-positive classification (CDR=1), whereas higher MMSE values indicate a lower likelihood of dementia.

**Fig. 5.**
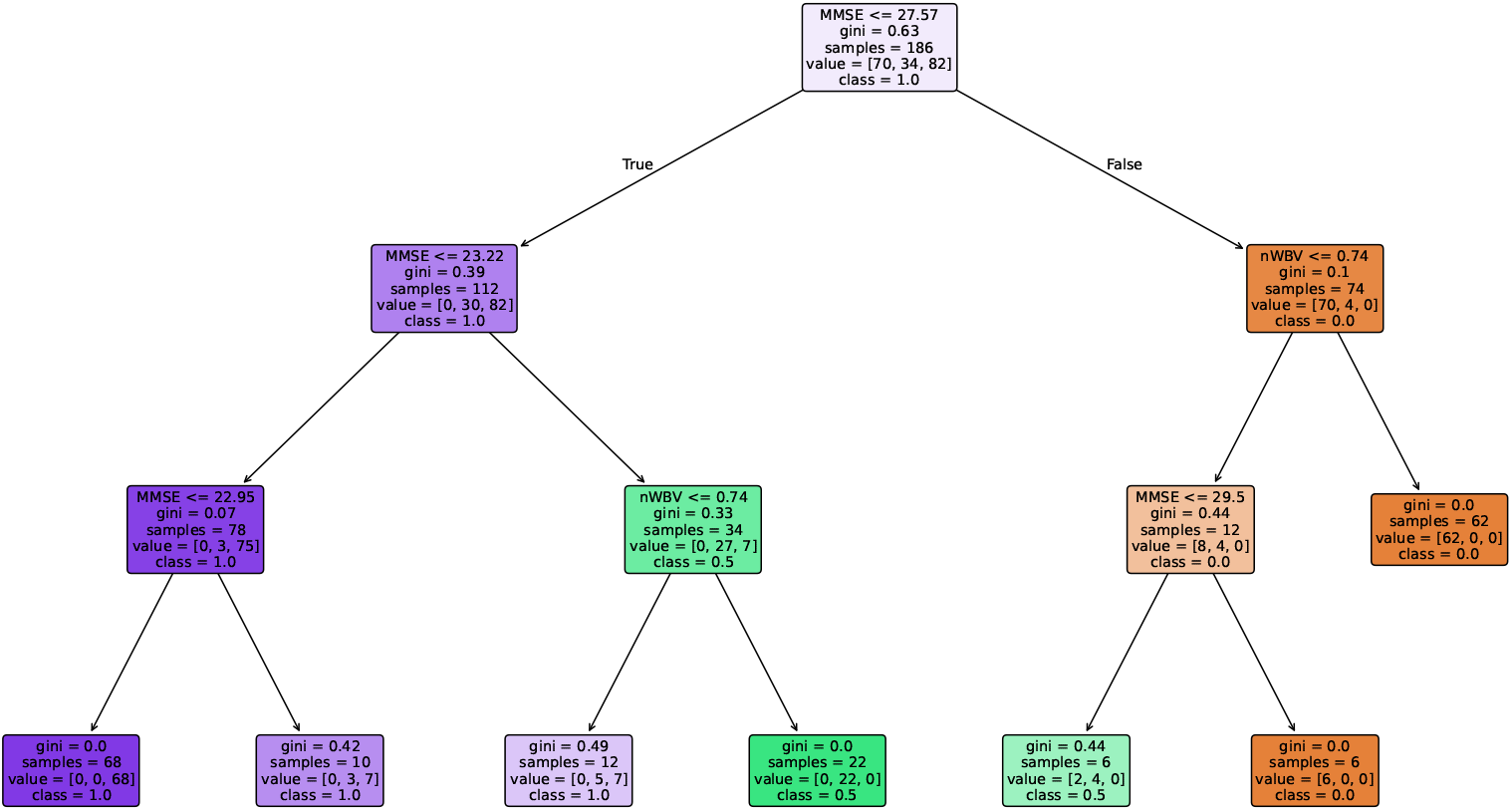
Decision tree constructed from training data with MMSE and nWBV features.

Within the subgroup of lower MMSE scores (MMSE *≤* 23.22), the tree further distinguishes cases by applying another MMSE threshold (MMSE *≤* 22.95). Subjects below this lower threshold demonstrate a strongly predominant dementia-positive classification (gini impurity = 0.07, classifying 75 out of 78 samples as dementia-positive). For subjects with MMSE values between 22.95 and 23.22, the decision tree incorporates nWBV as a secondary criterion. Specifically, subjects with lower normalized whole brain volume (nWBV *≤* 0.74) tend to be dementia-negative (class 0.5), indicating relatively preserved cognitive functions. Conversely, subjects with higher nWBV values within this subgroup are mostly classified as dementia-positive, albeit with higher uncertainty (gini impurity = 0.49).

The branch corresponding to higher MMSE values (MMSE *>* 27.57) leverages nWBV with a threshold of 0.74 . In this region, subjects with nWBV *≤* 0.74 show predominantly dementia-negative classifications, with very low uncertainty (gini impurity = 0.1). For higher nWBV values (nWBV *>* 0.74), the decision tree introduces an additional MMSE threshold (MMSE *≤* 29.5), differentiating subjects further into dementia-negative or positive. Notably, individuals with relatively lower MMSE values in this subset ( *≤* 29.5) predominantly belong to the dementia-negative group, though some uncertainty exists (gini impurity = 0.44). Individuals with even higher MMSE scores (*>* 29.5) unequivocally classify as dementia-negative, highlighting the predictive strength of MMSE at higher cognitive function levels.

Overall, the presented decision tree succinctly captures interactions between cognitive assessments (MMSE) and anatomical measures (nWBV). It offers clinicians transparent decision-making logic that directly connects feature thresholds to diagnostic outcomes, substantially enhancing clinical interpretability. The predictive performance of this interpretable model will be evaluated further on the validation subset, as detailed in the subsequent section. In the following section, we present detailed numerical experiments and comparative analyses to validate the effectiveness of our proposed method against existing state-of-the-art techniques.

## 4 Numerical Experiments

In this section, we empirically evaluate the predictive performance of our proposed explainable dementia detection method SADT against several benchmark machine learning algorithms commonly used with tabular clinical data. Specifically, we consider the following standard classifiers: Logistic Regression (LR), Linear Discriminant Analysis (LDA), Random Forest (RF), Support Vector Classifier (SVC), and Multi-layer Perceptron (MLP). Each of these models is evaluated under two experimental settings: first, using the original imbalanced data; second, employing oversampling via the Synthetic Minority Oversampling Technique (SMOTE) to address class imbalance.

Given the imbalanced nature of the dementia dataset, conventional metrics like accuracy alone may provide misleading insights. Therefore, we report evaluation results in terms of balanced accuracy and macro-averaged F1 score (F1 macro), which provide robust indicators of classification performance across both minority and majority classes. Each model is assessed through a 3-fold stratified cross-validation procedure to ensure consistent and unbiased performance estimation.

Table 2 summarizes the comparative results of our numerical experiments. The proposed SADT approach consistently outperforms all baseline models, both with and without oversampling, across both performance metrics. Specifically, SADT achieves a balanced accuracy of 0.667 *±* 0.024 and an F1 macro score of 0.638 *±* 0.042, surpassing all other methods evaluated. Among benchmark algorithms without oversampling, LDA and SVC present relatively competitive performances, achieving balanced accuracies of 0.637 *±* 0.073 and 0.630 *±* 0.100, respectively. However, their corresponding F1 scores are notably lower than SADT, indicating inferior handling of minority class predictions.

**Table 2.** Comparative evaluation of SADT versus benchmark ML methods.

|  | Balanced Accuracy | F1 Macro |
| --- | --- | --- |
| Logistic Regression | $0.614 \pm 0.090$ | $0.588 \pm 0.079$ |
| LDA | $0.637 \pm 0.073$ | $0.627 \pm 0.061$ |
| Random Forest | $0.599 \pm 0.072$ | $0.610 \pm 0.073$ |
| SVC | $0.630 \pm 0.100$ | $0.589 \pm 0.070$ |
| MLP | $0.578 \pm 0.076$ | $0.574 \pm 0.076$ |
| Logistic Regression-SM | $0.633 \pm 0.048$ | $0.584 \pm 0.024$ |
| LDA-SM | $0.616 \pm 0.088$ | $0.585 \pm 0.059$ |
| Random Forest-SM | $0.562 \pm 0.033$ | $0.558 \pm 0.030$ |
| SVC-SM | $0.649 \pm 0.083$ | $0.617 \pm 0.052$ |
| MLP-SM | $0.604 \pm 0.131$ | $0.575 \pm 0.098$ |
| **SADT (Proposed)** | **$0.667 \pm 0.024$** | **$0.638 \pm 0.042$** |

Interestingly, oversampling via SMOTE generally yields mixed results for the benchmark models. While oversampling slightly improves balanced accuracy for Logistic Regression and SVC, it adversely affects the Random Forest classifier, causing a noticeable decline in performance. Moreover, even the best-performing oversampled baseline model (SVC-SM) achieves a balanced accuracy of only 0.649 *±* 0.083, remaining substantially below the performance demonstrated by SADT.

These results strongly suggest that our integrated approach combining robust feature selection, advanced data balancing via SMOTE-ENN, and decision-tree-based interpretability offers distinct advantages. By directly addressing class imbalance while ensuring interpretability, SADT significantly enhances classification accuracy, providing a reliable, transparent, and clinically meaningful solution for dementia detection.

. These features of SADT may also make it a psychologically coherent explanatory framework. Future research should evaluate not only the model’s predictive power but also its decision-making and explanatory alignment with how clinicians conceptualize dementia categories and generate—and interpret—explanations. Considering how theories and principles from cognitive science can be integrated into the design and evaluation of AI systems could facilitate more robust and effective human-AI collaboration in clinical decision-making.

## 5 Conclusion

This paper introduced the Selectively Augmented Decision Tree (SADT), designed to address the dual requirements of predictive accuracy and clinical interpretability in dementia diagnosis. SADT integrates a three-phase pipeline consisting of feature selection using Random Forest importance and correlation analysis, advanced data balancing via SMOTE-ENN, and the construction of an inherently transparent decision tree classifier.

Our empirical evaluation, conducted on the OASIS dataset, demonstrated the effectiveness of the SADT approach. SADT achieved superior performance in terms of balanced accuracy and macro F1-score compared to several standard machine learning benchmarks, including Logistic Regression, LDA, Random Forest, SVC, and MLP, both with and without conventional SMOTE oversampling. These results validate the synergistic benefits of SADT’s integrated pipeline, highlighting the value of careful preprocessing combined with an interpretable modeling strategy. The resulting decision tree provides a clear, rule-based visualization of the classification process, identifying key diagnostic features (MMSE, nWBV) and their respective thresholds. The inherent transparency contrasts with “black-box” models, offering clinicians a tool whose reasoning can be directly inspected, understood, and verified against clinical knowledge.

Despite the promising findings, we acknowledge certain limitations. The study relied on a single, cross-sectional dataset (OASIS); validation on larger, longitudinal, and more diverse multi-center datasets would help ascertain the generalizability of SADT. The current model primarily addresses the presence of dementia based on CDR scores largely linked to Alzheimer’s Disease, and future work should explore its applicability to differentiating various dementia subtypes. In addition, while we propose a cognitive alignment, empirical studies are needed to directly evaluate how clinicians interact with and interpret SADT outputs in practice.

## Data Availability

All data produced are available online at: https://sites.wustl.edu/oasisbrains/home/oasis-1/

